# Biological Age and Age Acceleration Predict Alzheimer’s Disease Plasma Biomarker Levels

**DOI:** 10.1101/2025.05.22.25328181

**Authors:** Jaclyn M. Eissman, Yiyi Ma, Min Qiao, Dolly Reyes-Dumeyer, Angel Piriz, Annie J. Lee, Rafael A. Lantigua, Martin Medrano, Diones Rivera Mejia, Lawrence S. Honig, Francine Grodstein, David A. Bennett, Philip L. De Jager, Clifton L. Dalgard, Richard Mayeux, Badri N. Vardarajan

**Author notes:** Corresponding author Correspondence to: Badri N. Vardarajan, MS, PhD.

## Abstract

Epigenetic clocks can predict pathological aging associated with Alzheimer’s disease (AD) risk, albeit findings are mixed regarding if clocks are predictive in blood and in non-European populations. We constructed epigenetic clocks from blood methylation data in 704 older Hispanic adults and tested the association with a clinical diagnosis of AD and plasma biomarker levels. Biological age and age acceleration, the rate of biological aging, were significantly associated with sex, clinical diagnosis, and levels of eight plasma biomarkers, including P-Tau217 levels. Additionally, biomarker associations trended more significant among *APOE*-ε4 non-carriers. We also identified that methylation levels in CD4 and CD8 T-cell types are associated with biological aging and showed slightly stronger associations in men. We demonstrate that biological aging, in blood, in a Hispanic cohort of both demented and non-demented individuals, can stratify AD risk, predicting plasma biomarker levels even in preclinical disease.

## Main

Alzheimer’s disease (AD) is a public health crisis affecting 6.9 million Americans, and this number is expected to nearly double in the next 35 years^1^. Advanced age increases odds of developing AD, increasing 2-fold in every 10-year span from 65 to 85+ years old^1^. However, there is not a linear relationship between chronological age and AD risk, and there is much heterogeneity in aging trajectories over time. In fact, up to 30-50% of adults that receive a neuropathological classification of AD at autopsy were cognitively unimpaired throughout life^2^, demonstrating the need for novel biomarkers for early risk detection and stratification.

Epigenetic clocks were developed based on the premise that DNA methylation (DNAm) signatures mirror aging and disease at a biomolecular level^3^. Methylation patterns emerge with aging, including a global hypomethylation phenomenon resulting in “loss of transcriptional control” downstream and hypermethylation at CpG islands that are typically highly-methylated regions often found near promoters^4^. More specifically, studies have found changes at specific CpG sites that change with respect to aging, and these sites have been leveraged to calculate an individual’s “biological age”^5^. Brain DNAm changes have associated with AD neuropathology^6,7^, including in preclinical AD^6^, and accelerated epigenetic age has correlated with progression to MCI and AD as well as with cognitive decline^8^. Epigenetic clocks are also shown to accurately predict those that will go on to develop dementia^9,10^ up to 3 years prior to clinical diagnosis^10^. These studies support the vitality of epigenetic clocks as a potential early-stage AD biomarker.

Over the last decade, several epigenetic clocks of aging have been developed and validated, including Horvath^5^, Hannum^11^, and Levine (PhenoAge)^12^ clocks, which capture different aspects of aging. Additionally, cortical clocks^13,14^ were developed using postmortem brain methylation levels from the Religious Orders Study/Rush Memory and Aging Project (ROS/MAP)^13,14^. One limitation of most prior epigenetic clock studies is that they have been trained and applied to non-Hispanic whites, limiting their generalizability to more diverse groups. A recent study demonstrated that epigenetic clocks show a correlation with chronological age in all ancestry groups, but the associations were attenuated in individuals of non-European descent^15^. A second limitation is that, to our knowledge, biological aging associations with AD neuropathologic burden have primarily been established leveraging postmortem measures^7,13,16^. While these findings are quintessential to the validation of epigenetic clock measures with AD risk, there remains a need to establish whether epigenetic clocks can be successful tools for antemortem AD risk stratification.

Thus, in this study, we expanded on published work by calculating biological age and age acceleration in blood with two well-established aging clocks – Horvath and Hannum – in an aging Hispanic cohort. We evaluated the predictive power of these clocks for antemortem AD risk stratification, by testing associations with a panel of plasma AD biomarkers. We additionally examined associations stratified by sex and *APOE*-ε4 carrier status to have a more complete understanding of the association of biological aging with AD risk.

## Results

### Overview of study participants

Participants characteristics are outlined in **Table 1**. We leveraged 704 individuals from the Estudio Familiar de Influencia Genetica en Alzheimer (EFIGA) cohort study, including 169 clinical AD cases and 534 age-matched healthy controls. All individuals were of Caribbean Hispanic descent. Individuals with clinical AD had an older chronological age as compared to healthy controls and the full sample. AD was associated with higher biological age and faster age acceleration, on average, compared to healthy controls and the full sample. Our cohort included more women (65.05%) than men (33.81%) and contained 36.08% *APOE*-ε4 carriers (1 or 2 ε4 alleles) and 62.36% ε4 non-carriers. Additionally, in **Table 1**, we present biomarker averages and ranges among all individuals, healthy controls, and those with clinical AD. As expected, plasma P-tau 181 and 217 levels were higher in AD as compared to both healthy controls and the full sample, and the top end of the range of Aβ42/Aβ40 was slightly lower in AD cases. A clinical-pathological cohort of aging and AD, ROS/MAP, was leveraged for replication; please see **Methods** for more details on ROS/MAP.

**Table 1.**
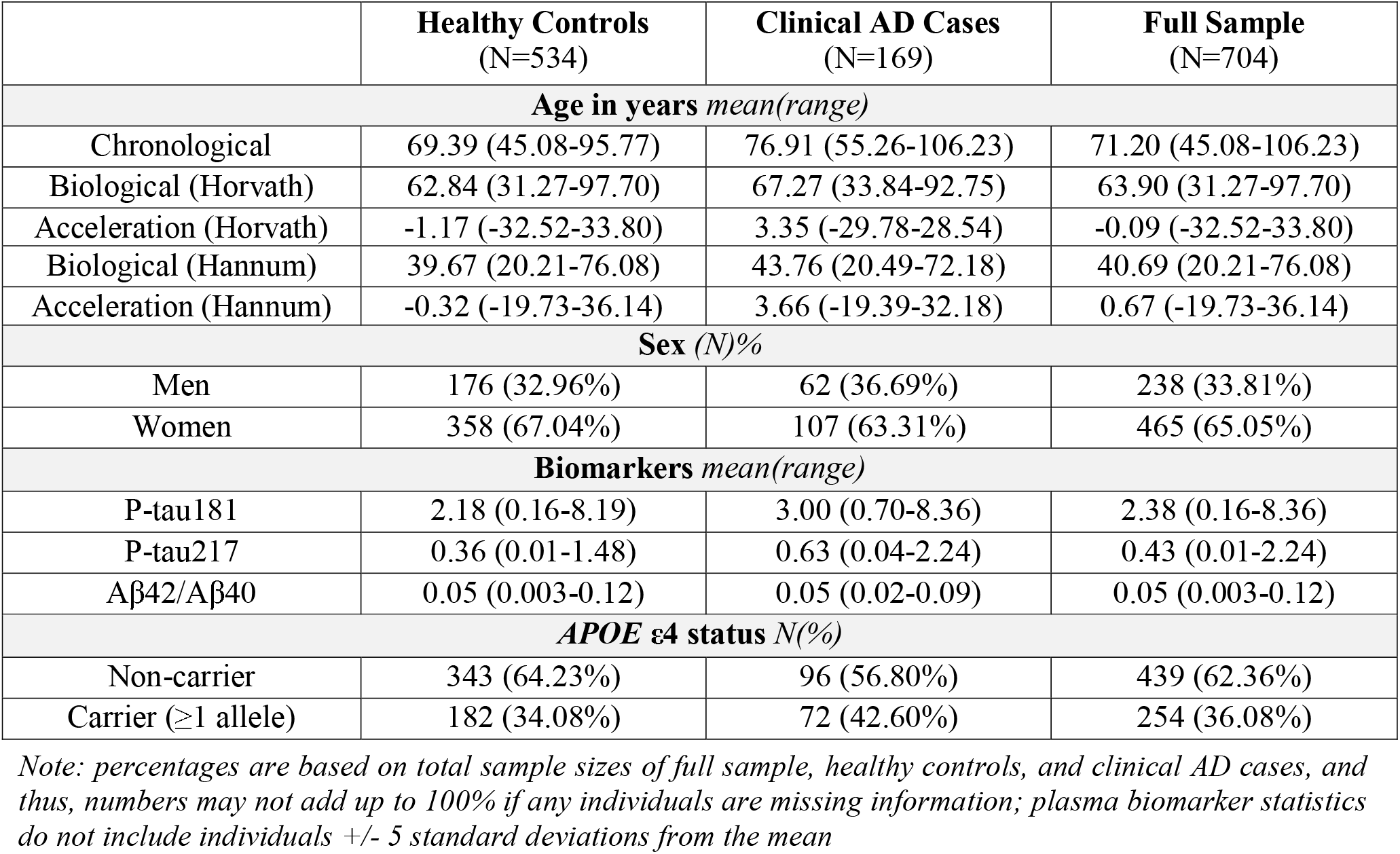
EFIGA Participant Overview.

|  | **Healthy Controls**<br>(N=534) | **Clinical AD Cases**<br>(N=169) | **Full Sample**<br>(N=704) |
| --- | --- | --- | --- |
| **Age in years** *mean(range)* |  |  |  |
| Chronological | 69.39 (45.08-95.77) | 76.91 (55.26-106.23) | 71.20 (45.08-106.23) |
| Biological (Horvath) | 62.84 (31.27-97.70) | 67.27 (33.84-92.75) | 63.90 (31.27-97.70) |
| Acceleration (Horvath) | -1.17 (-32.52-33.80) | 3.35 (-29.78-28.54) | -0.09 (-32.52-33.80) |
| Biological (Hannum) | 39.67 (20.21-76.08) | 43.76 (20.49-72.18) | 40.69 (20.21-76.08) |
| Acceleration (Hannum) | -0.32 (-19.73-36.14) | 3.66 (-19.39-32.18) | 0.67 (-19.73-36.14) |
| **Sex** (N)% |  |  |  |
| Men | 176 (32.96%) | 62 (36.69%) | 238 (33.81%) |
| Women | 358 (67.04%) | 107 (63.31%) | 465 (65.05%) |
| **Biomarkers** *mean(range)* |  |  |  |
| P-tau181 | 2.18 (0.16-8.19) | 3.00 (0.70-8.36) | 2.38 (0.16-8.36) |
| P-tau217 | 0.36 (0.01-1.48) | 0.63 (0.04-2.24) | 0.43 (0.01-2.24) |
| Aβ42/Aβ40 | 0.05 (0.003-0.12) | 0.05 (0.02-0.09) | 0.05 (0.003-0.12) |
| **APOE ε4 status** N(%) |  |  |  |
| Non-carrier | 343 (64.23%) | 96 (56.80%) | 439 (62.36%) |
| Carrier ($\geq 1$ allele) | 182 (34.08%) | 72 (42.60%) | 254 (36.08%) |

*Note: percentages are based on total sample sizes of full sample, healthy controls, and clinical AD cases, and thus, numbers may not add up to 100% if any individuals are missing information; plasma biomarker statistics do not include individuals +/- 5 standard deviations from the mean*

### Biological aging is associated with chronological aging

We first tested the association of chronological age with biological age, adjusted for sex. As expected, chronological age was significantly correlated with biological age (**Figure 1A-B)**, for both the Horvath clock (β=0.43, p=9.27×10^−33^) and the Hannum clock (β=0.41, p=1.45×10^−28^). We replicated these significant correlations leveraging the ROS/MAP brain epigenetic data, which confirmed significant correlations with chronological age for both the Horvath (β=0.71, p=3.90×10^−104^) and Hannum (β=0.60, p=5.41×10^−66^) clocks.

**Figure 1.**
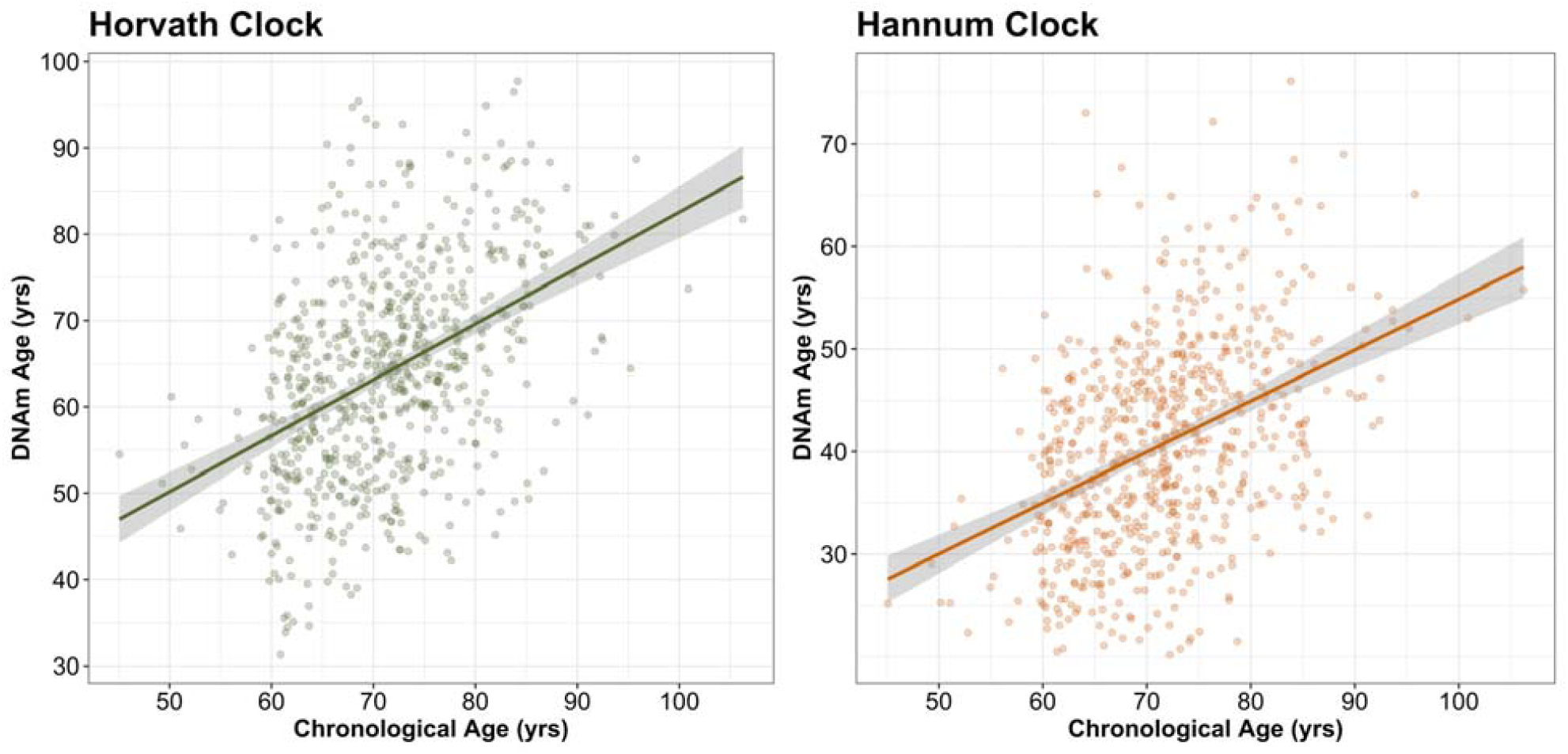
Biological age is significantly associated with chronological age. Biological age (DNAm) derived from A) Horvath and B) Hannum epigenetic aging clocks are significantly associated with chronological age in a Hispanic cohort of individuals with clinical Alzheimer’s disease and age-matched healthy controls.

### Biological aging is associated with sex

We tested the association of biological age and age acceleration with sex, adjusted for chronological age. Sex was significantly associated with biological age (**Figure 2A-B**), for both the Horvath clock (β=0.15, p=4.16×10^−2^) and the Hannum clock (β=0.17, p=1.88×10^−2^). Men had an older biological age and a faster biological age acceleration, on average, compared to women. We replicated these significant associations leveraging the ROS/MAP brain epigenetic data, which confirmed significant associations with sex for the Horvath clock (β=0.12, p=3.49×10^−2^) and the Hannum clock associated with sex but fell just below statistical significance (β=0.10, p=0.11). Age acceleration calculated from both the Horvath clock (β=0.14, p=4.83×10^−2^) and the Hannum clock (β=0.18, p=1.57×10^−2^) significantly associated with sex (**Figure 2C-D**). We additionally replicated these associations in ROS/MAP for the Horvath clock (β=0.12, p=3.25×10^−2^), but the Hannum clock fell just below the significance level (β=0.11, p=0.11).

**Figure 2.**
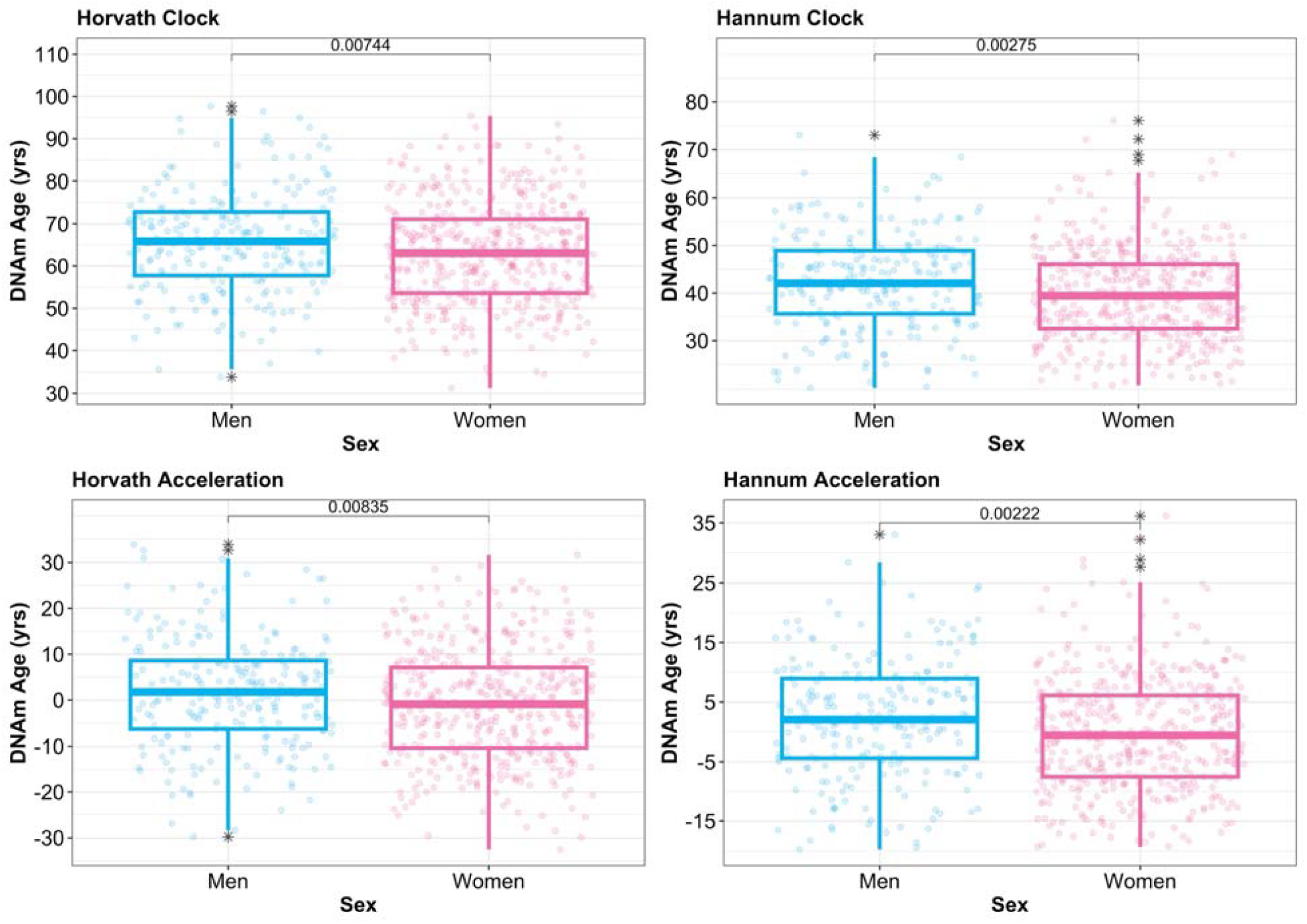
Biological aging is significantly associated with biological sex. Both biological age (DNAm) derived from A) Horvath and B) Hannum epigenetic clocks and age acceleration derived from C) Horvath and D) Hannum clocks are significantly associated with sex. Shown in panels A-D, men have both a slightly older biological age and a faster biological age acceleration as compared to women. P-values shown on the boxplots are derived from a simple t-test between sexes for biological age and age acceleration, respectively.

### Biological aging is associated with clinical diagnosis

We tested the association of biological age and age acceleration with clinical diagnosis (i.e., binarized variable: cognitively normal or clinical AD; **Figure 3A-B**), adjusted for sex. Clinical diagnosis (Horvath: β=0.36, p=4.67×10^−5^; Hannum: β=0.41, p=3.98×10^−6^) was significantly associated with biological age for both clocks. In ROS/MAP, we replicated the associations of biological age with clinical diagnosis (Horvath: β=0.51, p=7.93×10^−11^; Hannum: β=0.55, p=3.28×10^−12^). In addition, clinical diagnosis (Horvath: β=0.36, p=3.51×10^−5^; Hannum: β=0.40, p=7.66×10^−6^) was associated with age acceleration derived from both clocks (**Figure 3C-D**). We additionally replicated these associations with age acceleration in ROS/MAP (Horvath: β=0.51, p=5.97×10^−11^; Hannum: β=0.55, p=2.18×10^−12^).

**Figure 3.**
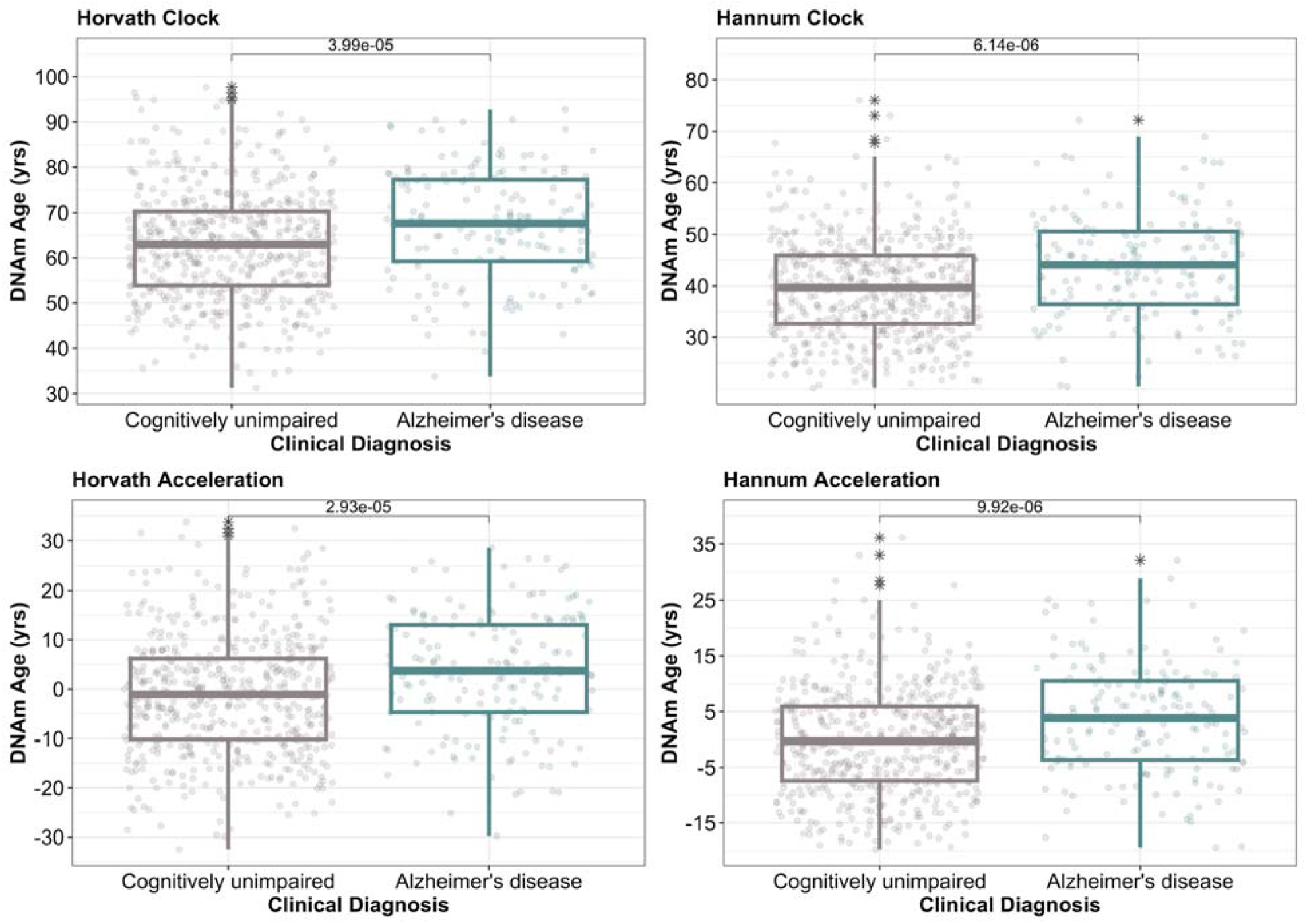
Biological aging is significantly associated with clinical diagnosis. Both biological age (DNAm) derived from A) Horvath and B) Hannum epigenetic clocks and age acceleration derived from C) Horvath and D) Hannum clocks are significantly associated with clinical diagnosis. Shown in panels A-D, those with a clinical Alzheimer’s disease diagnoses have both an older biological age and a faster biological age acceleration as compared to their aged-matched healthy counterparts. P-values shown on the boxplots are derived from a simple t-test between groups for biological age and age acceleration, respectively.

### Biological aging is associated with AD biomarkers and neuropathology

#### Biological Age

We tested the association of biological age with a panel of AD plasma biomarkers (**Figure 4A; Supplementary Tables 1,3,&5**) and considered associations significant if associated with at least one of the clocks (false discovery rate – FDR – <0.05). Of the 9 biomarkers tested, 8 were significant in at least one clock (**Figure 4A; Supplementary Table 1**), including P-Tau181, P-Tau217, and P-Tau231, Aβ42/Aβ40, Aβ40, GFAP, total-tau, and NfL, whereas Aβ42 was not associated in either clock. Top associations (considering both clocks) included P-Tau217 (Horvath: β=0.14, p=2.67×10^−4^, p.FDR=2.41×10^−3^; Hannum β=0.15, p=2.41×10^−4^, p.FDR=1.08×10^−3^) and NfL (Horvath: β=0.11, p=6.29×10^−3^, p.FDR=7.07×10^−3^; Hannum β=0.16, p=9.22×10^−5^, p.FDR=8.30×10^−4^). We validated biological age associations in ROS/MAP, leveraging autopsy neuropathology measures^17^, including the square root of the mean across 8 cortical regions for both amyloid plaque density (amyloid) and tau tangle density (tangles), a second summary measure of tau tangle burden (5 brain regions, silver-stained; NFT), as well as a global pathology summary score (i.e., neuritic plaques, diffuse plaques, neurofibrillary tangles; please see **Methods** for more details). In ROS/MAP, all 4 measures were significantly associated with biological age (**Supplementary Table 13**), including amyloid (Horvath: β=0.21, p=4.40×10^−8^, p.FDR=8.80^-8^; Hannum β=0.24, p=7.20×10^−11^, p.FDR=2.88×10^−10^) and tangles (Horvath: β=0.18, p=7.67×10^−7^, p.FDR=7.67×10^−7^; Hannum β=0.22, p=5.71×10^−9^, p.FDR=7.62×10^−9^). Notably, we observed a trend that biological age associations with biomarkers were more significantly associated among *APOE*-ε4 non-carriers as compared to ε4 carriers (**Supplementary Table 5**), and we likewise observed this pattern in ROS/MAP with the 4 autopsy measures of neuropathology (**Supplementary Table 15**). Please note, effect estimates of biological age on AD neuropathology in the ROS/MAP cohort are stronger than estimates observed in previous studies, possibly because we tested the effect of biological and chronological in separate models.

**Figure 4.**
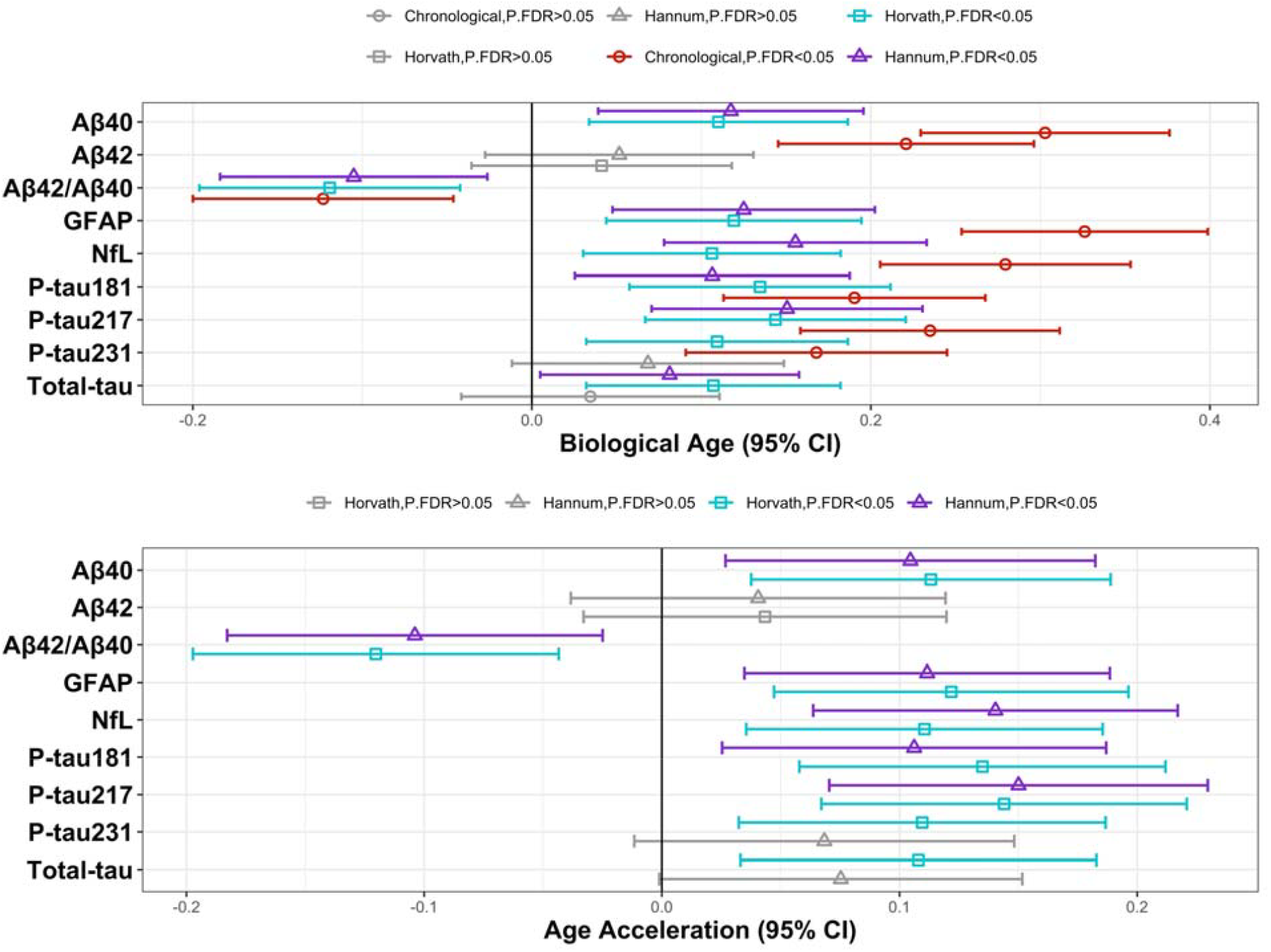
Biological aging is significantly associated with Alzheimer’s disease plasma biomarker levels. A) Biological age and B) age acceleration both derived from Horvath and Hannum epigenetic clocks are significantly associated with 8 out of 9 plasma biomarkers tested, with each biomarker listed on the x-axis. Both panels A and B show a 95% confidence interval calculated from the beta and standard error from each model. Age type is denoted by the shape and color of each plotted beta point estimate, with all nonsignificant results in grey.

#### Age Acceleration

We next tested the association of age acceleration with plasma AD biomarkers (**Figure 4B; Supplementary Tables 2,4,&6**) and considered associations significant if we identified significance with either clock (or both). Of the 9 biomarkers tested, 8 were significantly associated with age acceleration (**Figure 4B; Supplementary Table 2**), including all 3 P-tau measures 181, 217, and 231, Aβ42/Aβ40, Aβ40, GFAP, total-tau, and NfL, whereas Aβ42 was nonsignificant. Top associations (considering both clocks) included P-tau217 (Horvath: β=0.14, p=2.61×10^−4^, p.FDR=2.35×10^−3^; Hannum: β=0.15, p=2.40×10^−4^, p.FDR=1.62×10^−3^) and NfL (Horvath: β=0.11, p=4.00×10^−3^, p.FDR=5.99×10^−3^; Hannum: β=0.14, p=3.60×10^−4^, p.FDR=1.62×10^−3^). In our replication cohort (**Supplementary Table 13**), all 4 measures were significantly associated with age acceleration, including amyloid (Horvath: β=0.20, p=5.24×10^−8^, p.FDR=1.05×10^−7^; Hannum: β=0.24, p=9.07×10^−11^, p.FDR=3.63×10^−10^) and tangles (Horvath: β=0.18, p=9.77×10^−7^, p.FDR=9.77×10^−7^; Hannum: β=0.21, p=7.96×10^−9^, p.FDR=1.06×10^−8^). As with biological age, we observed a trend that age acceleration associations with biomarkers were more strongly associated among *APOE*-ε4 non-carriers as compared to ε4 carriers (**Supplementary Table 6**), and we likewise observed this pattern in ROS/MAP with the 4 autopsy measures of neuropathology (**Supplementary Table 15**).

### Chronological aging is associated with immune cell types

We deconvoluted the whole-genome methylation levels (please see **Methods**) to determine the cell-type proportional methylation levels in each participant. We then tested the association of each cell-type proportion with chronological age (**Figure 5; Supplementary Table 7**). Of the 12 cell-type proportions identified, 7 were significant after adjusting for multiple comparisons, including CD8 memory T-cells (β=0.13, p=1.12×10^−3^, p.FDR=8.29×10^−3^), naïve CD8 T-cells (β=-0.12, p=1.55×10^−3^, p.FDR=8.29×10^−3^), monocytes (β=0.12, p=2.07×10^−3^, p.FDR=8.29×10^−3^), CD4 memory T-cells (β=-0.11, p=3.51×10^−3^, p.FDR=1.05×10^−2^), naïve B-cells (β=-0.11, p=5.64×10^−3^, p.FDR=1.35×10^−2^), naïve CD4 T-cells (β=-0.10, p=7.08×10^−3^, p.FDR=1.42×10^−2^), and natural killer cells (β=9.21×10^−2^, p=1.53×10^−2^, p.FDR=2.62×10^−2^).

**Figure 5.**
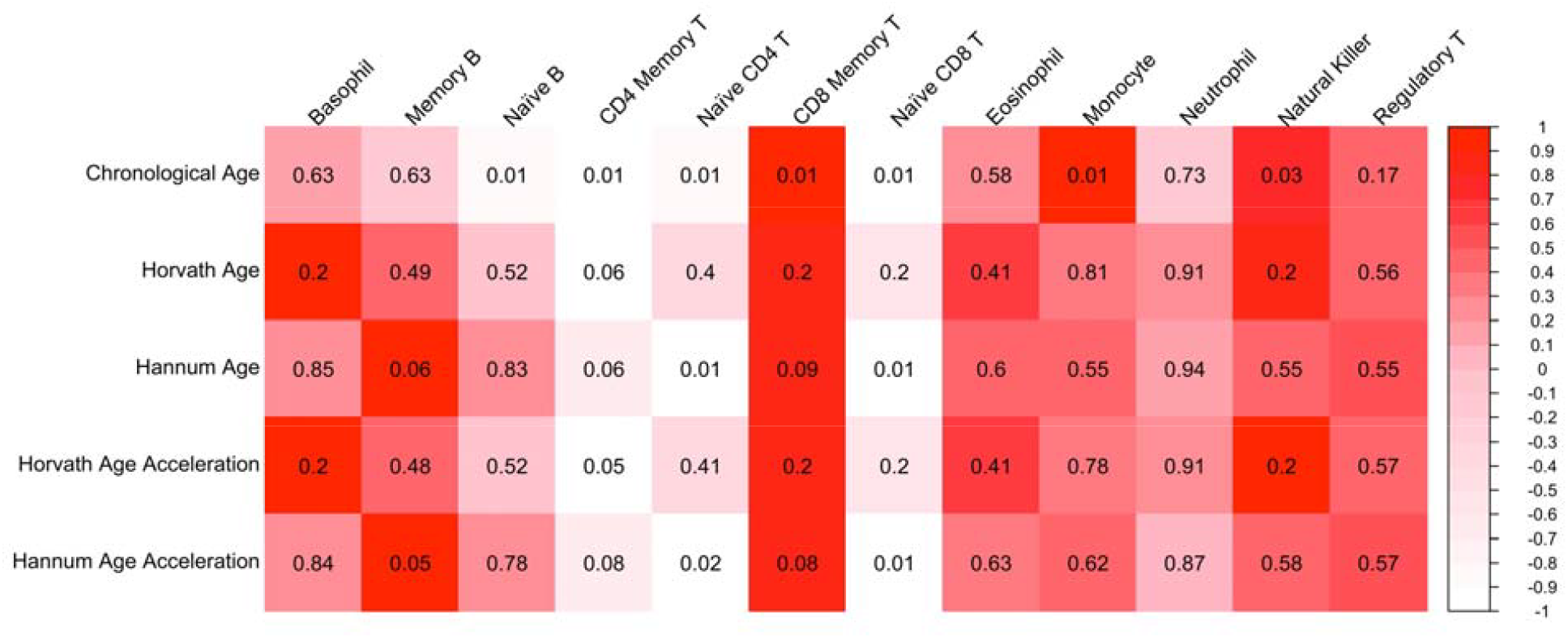
Biological aging is significantly associated with immune cell-type proportions. Red, white, and pink coloring represents strength of effect size, represented by beta values (re-scaled in figure from −1 to 1 for visualization purposes) from each model, and the black labeled values in each box represent the false-discovery-rate-adjusted p-values from each model. As shown, chronological aging is associated with multiple immune cell types (first row). Both biological age (second and third rows) and age acceleration (fourth and fifth rows) derived from Horvath and Hannum clocks show that CD4 and CD8 T-cells are significantly associated with both biological age and with rate of biological aging.

### Biological aging is associated with immune cell types

We assessed whether biological age (**Supplementary Tables 7,9,&11**) and age acceleration (**Supplementary Tables 8,10,&12**) were associated with cell-type proportions derived from deconvolution of the epigenetic data. Two cell types were significantly associated with biological age (derived from the Hannum clock; **Figure 5; Supplementary Table 7**), including naïve CD4 T-cells (Hannum: β=-0.12, p=2.46×10^−3^, p.FDR=1.48×10^−2^) and naïve CD8 T-cells (Hannum: β=-0.13, p=1.15×10^−3^, p.FDR=1.38×10^−2^). Additionally, the same two cell types, naïve CD4 T-cells (Hannum: β=-0.12, p=3.12×10^−3^, p.FDR=1.87×10^−2^) and naïve CD8 T-cells (Hannum: β=-0.13, p=1.23×10^−3^, p.FDR=1.48×10^−2^) were significantly associated with age acceleration (derived from the Hannum clock; **Figure 5; Supplementary Table 8**), with the addition of CD4 memory T-cells also reaching significance (Horvath clock-derived age acceleration; Horvath: β=-0.11, p=3.94×10^−3^, p.FDR=4.73×10^−2^). In addition, we found that both biological age and age acceleration (derived from the Hannum clock) were associated with three cell types more strongly in men compared to women (**Supplementary Tables 9-10**), including memory B-cells, naïve CD4 T-cells, and CD8 memory T-cells.

## Discussion

We conducted a comprehensive analysis of biological aging in Hispanics by building well-established epigenetic clocks in blood and testing associations with AD plasma biomarkers, replicating these findings in an independent cohort in the postmortem brain. First, we found that epigenetic age had weaker correlations with chronological age in Hispanics compared to previously published works in non-Hispanic white populations^13,15,16^. Next, we found that biological age acceleration was significantly associated with both clinical diagnosis and AD plasma biomarker levels in Hispanics, and that the associations with biomarkers were stronger among *APOE*-ε4 non-carriers compared to ε4 carriers. Epigenetic clocks are associated with AD and AD biomarkers, but their effect sizes are weaker than chronological age. This is probably because epigenetic clocks only capture a proportion of the variance in chronological age. Further, the rate of biological aging relates to the proportion of CD4 and CD8 T-cells, and this association trends stronger in men compared to women. Our study extends prior work on epigenetic clocks by demonstrating that biological aging can predict AD risk antemortem in a diverse sample that is enriched for preclinical AD.

The key finding of this study is that blood-based biological age and age acceleration relates to AD risk and AD plasma biomarker levels. We replicated our associations in an independent dataset in brain in ROS/MAP. Notably, prior literature about blood-based epigenetic clocks’ association with dementia risk has been inconsistent. While some studies show that blood-based epigenetic clocks could not predict risk^18–20^, other studies show that blood-based clock measures were associated with increased AD risk, the latter suggesting that these blood clocks are correlated with brain measures^7,12^. Our study helps to clarify this literature, showing that blood-based epigenetic clock measures are significantly associated with blood-based biomarkers, and we show that these associations replicate with brain epigenetic data and autopsy measures of AD neuropathology. This illustrates that biological aging and its associations with AD pathologic burden can all be measured antemortem and in a more noninvasive means than with lumbar puncture. This finding has implications for the possibility that blood-based epigenetic clocks could have clinical utility in the future in helping to stratify AD risk in patients, as epigenetic clocks address features of aging that chronological age does not and can predict an individual’s clinical and pathological AD risk.

We explored the relationship of biological aging with AD plasma biomarkers stratified by *APOE*-ε4 carrier status (the most robust known genetic risk factor for late-onset AD). Biomarker levels were more strongly associated with biological aging in ε4 non-carriers as compared to ε4 carriers. We interpret this to mean that biological aging may be a more important factor in AD pathogenesis among ε4 non-carriers, compared to ε4 carriers^21^. Biological aging may be one biochemical pathway that influences ε4 non-carriers’ risk for AD neuropathologic burden. We believe that biological aging may be a relevant pathway to stratify AD risk among ε4 non-carriers and that future work should evaluate specific DNA methylation site changes in ε4 non-carriers that may relate to AD risk and neuropathological burden among this group.

We found immune cell types were associated with both chronological aging and biological aging, whereby CD4 and CD8 T-cells were of relevance to biological aging. As expected, with increased chronological age, T-cell-related immunity changes included remodeling of T-cell composition^22^, which is a hallmark of aging. It is common to observe a decrease in naïve CD4 and CD8 T-cells and an increase in CD8 memory T-cells with age^23,24^, which we observed in our study, as well as observed an increase in proportions of both natural killer cell and monocyte composition and a decrease in naïve B-cell proportions with chronological age. When evaluating biological aging specifically, we observed a significant decrease in naïve CD4 T-cells, naïve CD8 T-cells, and memory CD4 T-cells, given older biological age and faster rate of age acceleration. This confirms prior evidence that slower biological age acceleration, or younger biological age^25^, is associated with higher naïve CD4 and CD8 T-cell composition^26,27^. Additionally, a recent study (in a diverse cohort) identified that “immunological age” was associated with entorhinal cortical thickness, an early region of change in AD, further emphasizing the importance of our findings in relation to AD risk^28^. Our sex-stratified follow-up analysis showed that T and B immune cell-type proportions appeared to be more relevant to biological aging in men as compared to women. One reason for this finding could be that most women in our study were post-menopausal, and menopause is considered an “inflection” point for immune system remodeling in women^29^. Possibly T- and B-cell composition was already lower in women^29^ in our study, and thus providing no evidence of change, whereas in men, this immune system change was detected in our study. Future studies will need to disentangle the relationship between sex, immunity, menopause, and biological aging.

Strengths of this study included patient stratification of AD using a less invasive measure of biological aging by leveraging plasma epigenetic data and plasma biomarker measures. Additionally, we were able to validate our biomarker associations in an independent cohort in the brain, showing that our findings are relevant to aging in the brain. Furthermore, we conducted this study in a Hispanic cohort, expanding our understanding of epigenetic clocks by providing evidence of its applicability across diverse groups. This study is not without limitations. The correlation of epigenetic clock measures with chronological age was lower in blood than in brain, which may have led to attenuated associations in our association testing with biomarkers. Additionally, as shown in other studies, biological age associations with chronological age are attenuated in non-European ancestry groups, possibly because the clocks are trained and constructed predominantly in non-Hispanic whites. Furthermore, cell-type composition relevant to aging in the periphery may differ from the composition relevant in brain aging, and cell types that did not have reliable markers for deconvolution may likewise be relevant to biological aging but could not be included in this study. Finally, our sample sizes of men/women and *APOE*-ε4 carriers/non-carriers were imbalanced, which may have led to some spurious associations and may have affected statistical power in the smaller groups.

We have demonstrated that plasma AD biomarker levels, directly related to AD neuropathologic burden, are associated with biological age and age acceleration in Hispanics, and that epigenetic changes may be a particularly relevant pathway for AD pathogenesis in *APOE*-ε4 non-carriers. We also show that a particularly relevant molecular pathway to rate of biological aging is “immunological aging,” specifically T-cell compositional changes. Our findings thus provide evidence for the future clinical utility of biological aging measures as an additional means for stratifying AD risk, while an individual is in preclinical disease stages.

## Methods

### Study participants

In this analysis, we leveraged the Estudio Familiar de Influencia Genetica en Alzheimer (EFIGA)^30^ cohort, which is a cohort study of aging and AD, comprised of older individuals with AD or aged-matched healthy controls with a family history of AD. All participants were of Caribbean Hispanic descent. Recruitment for EFIGA began in 1998 and took place in Washington Heights, New York and the Dominican Republic and participants were followed every 2 years. Participants had a family history interview, received an AD evaluation, neurological test battery, and medical and neurological exams. Additionally, NINCDS-ADRDA criteria^31,32^ was applied to determine if participants were cognitively unimpaired or had a probable or possible AD diagnosis, and the Clinical Dementia Rating^33–35^ was also used to help determine severity of disease. For this analysis, we created a binarized clinical diagnosis variable, whereby an individual was either classified as cognitively normal or clinical AD.

### Plasma biomarkers data generation

Blood for plasma was collected in dipotassium ethylenediaminetetraacetic acid tubes, and 2000g were centrifugated for 15 minutes at 4°C. The spun down blood was next aliquoted to polypropylene tubes, frozen, and stored at −80°C. Assays with specificity to each biomarker measured were applied to aliquots with the Simoa HD-X platform (Quanterix)^36^. Plasma biomarkers collected included Aβ40, Aβ42, total tau, GFAP, NfL, phosphorylated tau (P-tau) 181, 217, and 231, and we additionally calculated an Aβ42/Aβ40 ratio. For more detail on plasma biomarker data collection, please see our published protocol^36^. Prior to analysis, we removed values of biomarker levels that were more than 5 standard deviations above or below the mean across samples.

### Plasma epigenetic data generation

Blood was collected in PAXgene tubes at the same time as when the plasma biomarkers were collected as described above. To identify 5-methylcytosine and 5-hydroxymethylcytosine, we applied the NEBNext Enzymatic Methyl-Seq (EM-Seq) workflow^37^ with the following steps. First, 10ng of DNA was treated with TET2 oxidation of 5-methylcytosine, and second, modified cystines were deaminated with APOBEC. Next, PCR amplification was conducted with NEB Unique dual index primer pairs, and an Illumina NovaSeq 6000 sequencer was used for sequencing. Once sequenced, FASTQ files were aligned (GRCh38) leveraging the Bismark (v. 0.24.2)^38^ software package. Finally, duplicate reads that had MAPQ scores <20 were subsequently filtered out. We additionally performed a de-convolution step on the cleaned whole-genome epigenetic data with the EpiDISH^39^ package in R. Our de-convoluted data contained the following cell-type proportions for which we had reliable markers: CD4 and CD8 memory T-cells, naïve CD4 and CD8 T-cells, regulatory T-cells, natural killer cells, naïve and memory B-cells, eosinophil, basophils, monocytes, and neutrophils.

### Building biological age and age acceleration

We built epigenetic clock ages (i.e., biological age) with the “methylclock” R package^40^ that is publicly available on GitHub (https://github.com/isglobal-brge/methylclock). To prepare for clock calculations, we first matched chromosome and base pair positions from our methylation data with the Illumina Array Infinium Methylation EPIC (v. 2.0) CpG Product File annotations (GRCh38). This was necessary as the clocks each used Illumina array CpG site nomenclature. Before clock calculations, we filtered out a sample or a CpG site if it had >20% missing data. We calculated 2 clocks with the *DNAmAge* function (methylclock package), which allows for calculation of the Horvath^5^ and Hannum clocks^11^, both of which leverage elastic net modeling. Any missing data at a required CpG site were imputed during clock age calculations, which by default of the methylclock packages, implements the *impute*.*knn* function (impute R package), applying the K-nearest neighbor method for imputation^40^. In addition to calculating biological age, we calculated age acceleration with the methylclock package, which is defined by the residual of biological age regressed onto chronological age.

### Association testing with biological age and age acceleration

All association testing was conducted in R with linear models for quantitative outcomes and generalized linear models for binary outcomes. Prior to association testing, biological age and age acceleration outlier values were removed based on visual inspection of histograms. Biological age and age acceleration from each clock served as predictors for the models. If quantitative predictors and outcomes were not already scaled, we scaled (default scale function in R) variables in the model to get standardized output. All models were adjusted for self-reported sex. We tested the effect of biological age without adjustment of chronological to avoid collinearity in the regression models. We present the association results of both chronological and biological age from independent models. To test the residual effect of biological age after adjustment for the effect of chronological age, models with age acceleration (as the predictor) were used. An identical set of models were performed stratified first by sex (note – these models did not covary for sex) and then by *APOE*-ε4 carrier status (defined by presence of 1-2 ε4 alleles vs. no ε4 alleles). Within each set of outcomes, the false-discovery rate (FDR) procedure was used to adjust p-values within each analysis, with significance set at FDR<0.05. FDR-correction was performed separately per clock, and we considered an association significant if an outcome survived FDR-correction in one or both clocks.

### Replication of biological aging findings

We sought replication of our findings in an independent cohort, the Religious Orders Study (ROS)/ Rush Memory and Aging Project (MAP), a well-established aging and AD cohort^41^. ROS and MAP are both longitudinal clinical-pathological studies that followed participants from the Chicago area over time, whereby ROS recruited nuns, priests, and brothers from religious orders, and MAP recruited community-dwelling participants from retirement and senior living areas. ROS/MAP participants enroll without known dementia and agree to annual neurological exams and cognitive performance tests and brain donation upon death. Both studies were approved by an Institutional Review Board of the Rush University Medical Center. All participants signed an Anatomic Gift Act and informed and repository consents. For generalization in ROS/MAP, we calculated biological age and age acceleration leveraging identical methods as described above. Further, to maximize sample size, we retained samples of both Hispanic and non-Hispanic descent, and of all ancestry groups. To replicate biomarker associations in brain, we tested biological age and age acceleration associations with autopsy measures of neuropathology^17^, including 4 measures: the square root of the mean for 8 quantified measures of (1) cortical amyloid plaques (amyloid; quantified by immunohistochemistry) and (2) neurofibrillary tau tangle density (tangles; AT8, immunohistochemistry), (3) mean count of tau tangles divided by standard deviation of 5 silver-stained brain regions (NFT), as well as (4) a quantitative summary score for neuritic and diffuse plaques and neurofibrillary tangles across 5 brain regions (global pathology). We also created a binarized clinical diagnosis variable, whereby individuals with no cognitive impairment or mild cognitive impairment were in one group and those with clinical AD were in another group.

## Acknowledgements

Data collection for this project was supported by the Genetic Studies of Alzheimer’s disease in Caribbean Hispanics (EFIGA) funded by the National Institute on Aging (NIA) and by the National Institutes of Health (NIH) (5R37AG015473, and R01AG067501). ROS/MAP is supported by P30AG10161, P30AG72975, R01AG15819, R01AG17917, U01AG46152, and U01AG61356.

## Data availability

All results are included in the main text or the supplementary materials of the manuscript. The raw plasma biomarker and whole-genome methylation data will be shared with qualified investigators using the request form available here: https://cumc.co1.qualtrics.com/jfe/form/SV_dmck0uV3A91pmzb. ROS/MAP resources can be requested at www.radc.rush.edu and http://www.radc.rush.edu and www.synpase.org.

## Code availability

Code from this study will be made available on GitHub: https://github.com/jaclyn-eissman.

## Author information

### Author contributions

Conceptualization: JME, BNV

Methodology: JME, BNV, CLD

Participant enrollment and sample collection: DRD, MM, DRM, RAL, LSH, RM

Data Generation: MQ, AP, CLD, AJL, LSH, DAB, PLDJ, RM, BNV

Data analysis and interpretation: JME, BNV

Funding acquisition: RM, BNV

Project administration: DRD, RAL, RM

Supervision: BNV

Writing: original draft: JME, BNV

Writing: reviewing & editing: all authors

